# From Adversity to Pain: Disrupted Insulo-Cingulo-Thalamic Dynamics in Emotional Abuse and Neglect

**DOI:** 10.1101/2025.09.23.25336443

**Authors:** Antoniou Georgia, Smith H. Blair, Steele J. Douglas, Colvin A. Lesley

## Abstract

Adverse Childhood Experiences (ACEs) increase vulnerability to chronic pain and mood disorders, yet the neural mechanisms linking early adversity to pain persistence remain poorly understood. We investigated whether ACEs are associated with altered effective connectivity within the salience network, a system critical for integrating emotionally and physiologically salient information and strongly implicated in chronic pain. Using neuroimaging data from a well-characterised, longitudinal cohort, participants performed an emotion-processing task (fearful vs. neutral faces) during fMRI. Region-of-interest analyses and dynamic causal modelling (DCM) were applied to examine directional connectivity among the anterior cingulate cortex (ACC), insula, and thalamus in relation to self-reported ACEs and chronic pain. Chronic pain and childhood emotional neglect were associated with reduced ACC-to-insula connectivity, suggesting impaired top-down regulation of interoceptive and affective signals. In contrast, individuals with chronic pain and a history of emotional neglect or abuse showed increased thalamus-to-insula and thalamus-to-ACC connectivity, consistent with heightened salience of sensory and emotional inputs. These findings demonstrate that ACEs influence salience network dynamics, establishing a mechanistic link between early adversity and chronic pain. They suggest that ACEs modify brain network activity, potentially contributing to the persistence of chronic pain and emphasising the roles of the ACC, insula, and thalamus.

## Introduction

Adverse Childhood Experiences (ACEs) are events or situations likely to cause harm or distress that a child experiences or encounters within their environment, undermining their sense of safety[62,85]. These experiences can include abuse (such as physical, sexual, or emotional abuse), neglect (such as physical or emotional neglect), poverty, household dysfunction (domestic violence or substance abuse), or the loss of a parent[18,46,57,62,85]. Exposure to ACEs is associated with a broad range of adverse outcomes, including psychiatric disorders, chronic disease, and premature mortality[19,53]. Multiple studies also link ACEs to an increased risk of chronic pain[12,51], a condition that represents the leading cause of disability worldwide[41]. In the UK, chronic pain affects over one-third of adults and frequently co-occurs with depression[15,20,45,60], underscoring the need to clarify mechanisms that connect early adversity to pain vulnerability.

ACEs can alter brain development at the epigenetic, cellular, and network levels[34,47,55]. Despite the well-established links between ACEs, chronic pain, and depression, the neurobiological mechanisms underlying these associations remain poorly understood. While some neuroimaging studies have investigated these relationships without the comorbidity, none have examined them concurrently within the same population, particularly in a large community-based sample[3].

Neuroimaging studies suggest that ACEs alter regions implicated in emotion and pain processing[56]. For instance, exposure to childhood violence has been associated with heightened insula activation[16,28,52,54,58,59], whereas survivors of assault show reduced activation in the anterior insula and anterior cingulate cortex (ACC), reflecting disrupted trust and threat processing[48]. ACEs have further been associated with reduced grey matter volume in the ACC[17] and altered responses to threat cues[42,89]. Despite these insights, no studies have examined how ACEs and chronic pain jointly influence connectivity within core pain-related networks in a large, community-based sample.

The insula, ACC, and thalamus are central to the salience network, which integrates sensory, affective, and interoceptive information to guide adaptive responses. The anterior insula translates nociceptive input into the subjective experience of pain[7,9,11,29,80] and has been proposed as a robust biomarker for chronic pain[21]. The ACC contributes to both affective and cognitive aspects of pain, as well as emotion regulation and threat detection. At the same time, the thalamus serves as a relay hub transmitting nociceptive signals to cortical regions, including the insula and ACC[2,14,23,44,65]. Aberrant functioning within this insulo-thalamo-cingulate circuit may therefore represent a neural mechanism through which ACEs increase vulnerability to persistent pain.

Based on this framework, we hypothesised: (1) that insulo-thalamo-cingulate connectivity would be associated with chronic pain severity, reflecting altered integration of sensory and affective pain signals[35,40]; and (2) that these alterations would be most pronounced in individuals with both chronic pain and a history of emotional adversity, specifically childhood emotional abuse and neglect. By examining insulo-thalamo-cingulate connectivity in individuals with chronic pain and ACEs, we aimed to identify network alterations that may underlie the persistence of adversity-related pain and highlight potential neural targets for intervention.

## Methods

### Participants

Data from the Generation Scotland Scottish Family Health Study (GS:SFHS) and the subsequent Stratifying Resilience and Depression Longitudinally (STRADL) study was used in this study. The GS:SFHS dataset of around 24,000 community-based participants (aged over 18) contains detailed socio-demographic and clinical data collected at study entry between 2006-2011[78]. A subset of the GS:SFHS participants took part in the subsequent STRADL study 3-11 years later[36,64]. Participants were invited to complete various face-to-face assessments, sample collection and brain magnetic resonance imaging (MRI) scanning either at Aberdeen or Dundee Universities.

### Public and patient involvement

The Chronic Pain Advisory Group (CPAG), comprising individuals with lived experience of chronic pain and ACEs, contributed to the development and refinement of the research questions. Members of the CPAG were involved throughout the research process as part of the Consortium Against Pain inEquality (CAPE), which prioritises inclusive and participatory research practices.

### Questionnaire data and clinical interviews

All participants were assessed for a lifetime history of MDD. Repeating the GS:SFHS baseline assessment of depression in the STRADL study, a research version of the Structured Clinical Interview for DSM-IV disorders (SCID)[22] was used to assess symptoms of mood disorder (including MDD and episodes of mania and hypomania) according to the Diagnostic and Statistical Manual of Mental Disorders (DSM-IV-TR) criteria. For the assessment of the severity of depressive symptoms, the Quick Inventory of Depressive Symptomatology (QIDS)[43] was administered, a 16-item mood questionnaire. Adverse childhood experiences (childhood or adolescent abuse or neglect) were assessed using the Childhood Trauma Questionnaire short form (CTQ-SF)[8]. This is a 28-item quantitative and retrospective inventory that assesses histories of abuse and neglect; the subscale of the questionnaire includes emotional, physical, and sexual abuse and emotional and physical neglect

Chronic pain was assessed through a pre-clinic questionnaire in the GS:SFHS study, which occurred about a decade earlier than the STRADL study. Participants were asked if they experienced continuous or intermittent pain, and if yes, whether this pain had lasted for at least 3 months or more. Those answering yes to both questions, the Chronic Pain Definition (CPD) questionnaire[70], were classified as having chronic pain and were asked to complete additional questions to assess chronic pain severity using the CPG questionnaire[79,87]. This seven-item questionnaire classifies respondents into four grades of severity: CPG 1 (low disability, low intensity), CPG 2 (low disability, high intensity), CPG 3 (high disability, moderately limiting) and CPG 4 (high disability, severely limiting). Odds ratios for associations with socio-demographic variables were calculated.

### fMRI data acquisition

T1-weighted images and functional MRI data were acquired at Dundee and Aberdeen Universities. In Aberdeen, participants were imaged on a 3T Philips Achieva TX-series MRI system (Philips Page 10 of 31 Healthcare, Best, Netherlands) with a 32-channel phased-array head coil and a back facing mirror (software version 5.1.7; gradients with maximum amplitude 80 mT/m and maxi-mum slew rate 100 T/m/s). For the functional MRI acquisition in Aberdeen, a projector and “Presentation” (Neurobehavioural Systems Inc, Berkeley, CA, USA) version 18.1 were used with a repetition time of 1.56 s, echo time of 26 ms, flip angle of 70^◦^, field of view 217 mm, matrix 64×64, 32 axial slices were used. In Dundee, participants were scanned using a Siemens 3T Prisma-FIT (Siemens, Erlangen, Germany) with 20 20-channel head and neck phased array coil and a back-facing mirror (Syngo E11, gradient with max amplitude 80 mT/m and maximum slew rate 200 T/m/s). A magnetic resonance compatible LCD screen was used to display fMRI (NordicNeuroLab, Bergen, Norway) task stimuli using “Presentation” version 20.0 using repetition time 1.56 s, echo time 22 ms, flip angle 70^◦^, field of view 217 mm, matrix 64×64 and 32 axial slices.

A blocked fMRI design was used to analyse the implicit emotional processing task with 6 blocks. A particular focus was to assess emotional-limbic circuitry and the neural responses to viewing fearful faces. The participants observed a block of 6 faces (neutral or fearful); see **Figure 1**. To avoid a gender bias with the images, two versions of the tasks were used, counterbalanced across participants. The NimStim facial expression set was used to display neutral and fearful faces[83].

**Figure 1:**
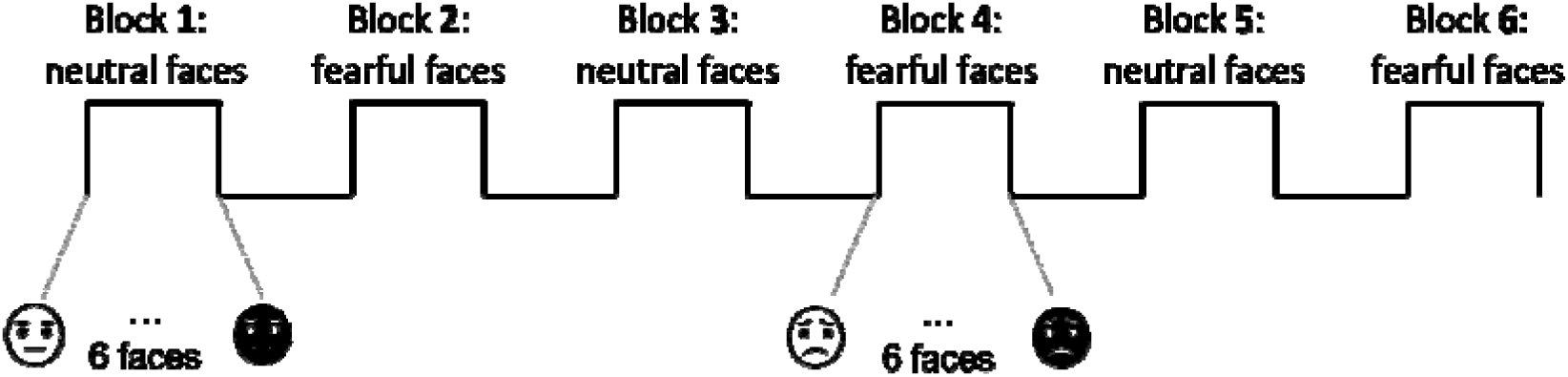
Implicit emotional processing task (fearful versus neutral faces): block fMRI design, 6 blocks of neutral and fearful faces from NimStim set of facial expressions set. Two versions of the task were used in and counterbalance across participants, to avoid a gender bias of the images.

### Image Pre-processing and First-level Analysis

The functional MRI images were analysed using Statistical Parametric Mapping 12 (SPM12), and a manual check for artefacts was conducted prior to the pre-processing analysis, and the first eight blood oxygen level-dependent volumes were discarded as standard because of transient effects. Functional images were realigned to match the first image by applying a six-parameter rigid body transformation; then the T1-weighted structural image was co-registered to the realigned functional images. The T1-weighted structural image was then segmented into different tissue types using the SPM12 tissue probability maps, and the resulting deformation field was used to normalise the images. The final step of the pre-processing analysis involved using an 8 mm full width at half maximum Gaussian kernel to smooth the functional images.

An event-related design was used for the first-level analysis. A first-level general linear model (GLM) design matrix included two columns of possible outcomes: fearful and neutral faces. The six rigid body motion realignment parameters estimates obtained during pre-processing were included as covariates of no interest. Events were modelled as truncated delta functions, which were then convolved with the SPM12 canonical haemodynamic response function without time or dispersion derivatives.

Random effects, event-related fMRI analyses were done using data from 579 participants, which included 238 subjects with chronic pain, 60 subjects with emotional neglect and 79 subjects with emotional abuse. As recommendations for DCM[26,91,92], data from 371 subjects who had sufficiently strong functional MRI signals in the regions of interest for analyses were selected.

### Dynamic Causal Modelling (DCM) Analysis

The DCM, as implemented in SPM12, was used to analyse the effect of the emotional processing task on effective connectivity within brain regions, exploring whether the effective connectivity strengths were related to reported chronic pain and ACEs. DCM allows the investigation of the relationships between inputs, internal states, and outputs across a network of brain regions[25]. The inputs are external stimulus functions (task variables), internal states correspond to neuronal and neurophysiological variables necessary to produce outputs, and the outputs are the hemodynamic responses measured by fMRI. Inputs can elicit responses in two ways. One way is by directly activating specific brain regions, for example, a visual stimulus may directly activate the visual cortices. The second is by indirectly influencing the connections among brain regions by modulating the coupling among nodes. In the DCM method, the interactions between brain regions when no specific task is being performed are referred to as intrinsic connections, and coupling between nodes caused by experimental conditions is referred to as modulation.

The driving input is typically represented as the immediate impact of experimental stimulus information on specific brain regions. In the emotional processing task, fearful and neutral faces were used, so the visual stimuli directly activated occipital regions, subsequently driving inputs to the rest of the brain network examined in this study.

### Time series extraction

Brain volumes of interest (VOI) were selected to test the hypotheses of chronic pain and ACEs changes in effective connectivity between regions which are part of the salience and pain processing networks. The current study identified the coordinates of the left ACC, insula, and thalamus by using regions of the brain that showed significant activity when participants viewed fearful faces compared to neutral ones. The time series of VOIs were extracted from second-level peak coordinates by creating masks using WFUPickAtlas for the ACC, insula and thalamus, allowing SPM to find peak coordinates within the masks for each individual. The MNI coordinates of the VOIs were surrounded with 8mm spheres, and the first principal component of the time series was extracted for each participant.

The selection of the left ACC, Insula, and Thalamus was based on their established role in emotional regulation and stress response mechanisms[30,32,69,76,81,88], particularly in relation to the dysfunction and structural changes that appear in individuals with ACEs[5,38,49,82,90] and chronic pain[4,10,27,33,40,93]. The Thalamus, in addition to its role in processing sensory information such as pain[35,40] has extensive connections to various other brain regions, much like the ACC and Insula[2,14,23,31,65,76]. The anterior insula and anterior cingulate are linked to the experience of pain but are also activated by other aversive experiences, including negative affect and anxiety[66,76] and the insula and ACC have reciprocal projections[14,23,31]. The spinothalamic tract transmits information about pain to the posterior thalamus, with its main cortical projections being the posterior insula, parietal operculum, and mid-cingulate cortex[29].

### Bayesian model selection and averaging

A DCM model with bi-linear dynamics and a single state per region was assumed with no stochastic effect and mean-centred. A fully connected model with nine connections, including self-inhibitory connections, was fitted in the first-level analysis, and the percentage of the variance that was explained was calculated. The PEB (Parametric Empirical Bayes) framework[26,91,92] was employed to model similarities and variations among participants. At the second level, individual DCMs underwent Bayesian Model Reduction (BMR) to eliminate connections by estimating different reduced PEB models within which certain parameters were ‘switched off’[92]. A “greedy” automated search procedure was employed to iteratively eliminate parameters that had no impact on the free energy[92]. For statistical comparison of the model parameters, Bayesian model averaging (BMA) was performed to compute the average of DCM parameter estimates across the entire model space, with weights based on the posterior probabilities of each model[67,91,92]. The PEB design matrix at the group level for between-subject comparison included a column of ones representing the average connectivity across participants and a zero-mean centred column of the covariates of interest, such as reported chronic pain, emotional abuse, and emotional neglect scores. The within-subject design matrix at the group level was specified as the identity matrix, implying that the covariates could potentially affect each within-subject DCM parameter. The complete PEB model was inverted to calculate parameter estimates and the “free energy” of the model.

## Results

### Participants

Event-related functional MRI analyses were performed on 579 participants, which included 238 subjects with chronic pain, 60 subjects with emotional neglect and 79 subjects with emotional abuse. For DCM, 371 subjects with sufficiently strong functional MRI signals were selected in the regions of interest. Demographic details of the population are presented in **Table 1**.

**Table 1:**
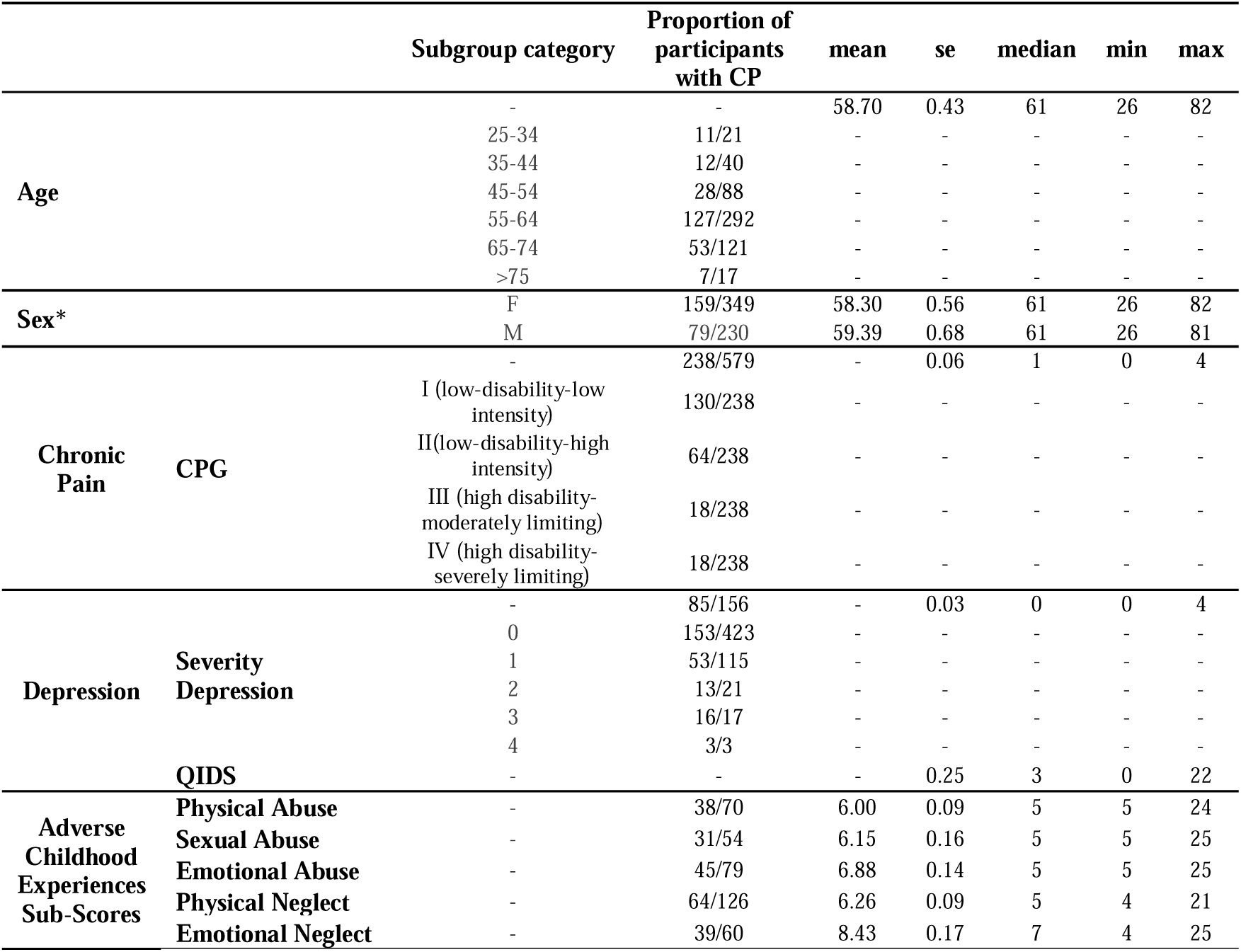
Clinical and demographic characteristics of the population. Includes associations of age, sex, depression, and adverse childhood experiences with ‘chronic pain’. Age was assessed during face-to-face entry of the STRADL study. Quick Inventory of Depressive Symptomatology; QIDS, chronic pain grade; CPG, adverse childhood experiences; ACEs.

The odds of having depression co-morbid with chronic pain were found to be 2.11 (95% CI 1.46 to 3.07) times greater than the odds of having depression without having chronic pain. Moreover, the odds of having chronic pain for people exposed in childhood to emotional abuse were 2.11 (95% CI 1.30 to 3.40) times greater than the odds of having chronic pain for participants who were not exposed in childhood to emotional abuse, whereas for physical abuse the odds were 1.97 (95% CI 1.18 to 3.28) times greater, for sexual abuse were 2.07 (95% CI 1.17 to 3.65), for physical neglect were 1.66 (95% CI 1.11 to 2.46) times greater, and for emotional neglect, they were 2.99 (95% CI 1.71 to 5.22) times greater than those who were not exposed to the respective adversity in their childhood.

### General linear model voxel-based functional MRI analyses

As expected, across all 579 participants, significant de-activations regarding fearful and neutral faces were identified in areas including the amygdala/ hippocampus (−32 −26 −10), the medial superior frontal cortex (8 38 24), ACC (−8 46 14) (12 38 2), the mid-cingulate (40 26 42), the anterior insula (−30 16 6), thalamus (2 −18 16), as well as deactivations in the caudate/putamen (22 24 12). Details are provided in the supplementary material. For the chronic pain participants, as hypothesised, negative correlations between fearful faces activation magnitude and CPG scores, chronic pain severity scores, were found in areas that include the nucleus accumbens (−6 12 −12), parahippocampal (−22 −28 −22), superior frontal (−16 20 66) (22 26 34), Heschl (38 −24 12) gyri. A positive correlation with CPG scores was found in the superior motor area (10 4 18), mid-cingulate (16 −22 50) and putamen (−28 4 −8).

For participants who had experience emotional neglect in childhood, negative correlations between fearful faces activation magnitude and SA sub-scores of the CTQ were found in areas that include the pallidum (14 −30 22) (20 8 0), thalamus (−14 −8 0) (12 −30 16), posterior and anterior cingulate cortex (18 54 12) (−14 −36 20), caudate and superior and middle frontal (−20 60 10) (−28 46 10) gyri. A positive correlation with CPG scores was found in the amygdala (−22 0 −22) (−42 10 −30), superior frontal (18 10 44), superior motor area (−10 4 54), as well as activations in the occipital lobe visual areas (10 −88 12).

### Dynamic causal modelling of event-related effective connectivity

The mean of the explained variance across the 371 participants with sufficiently strong functional MRI signals in the regions of interest was 19.21%. There were no statistically significant differences in mean covariates scores (QIDS, CTQ, Total EA, Total EN, Total SA, Total PA and Total PN) between the 371 participants with explained variance *>*10% and the total of 578 participants.

Nearly all the connections showed strong evidence in the DCM analysis, based on the criterion of free energy (p*>*0.95), except the connection from the ACC to insula and from thalamus to insula in terms of effective connectivity, see Figure 2(a). The second level PEB analysis revealed that chronic pain patients’ effective connectivity from insula to thalamus increased. Additionally, insula to ACC connection had a negative association in chronic pain patients who had experienced emotional neglect in their childhood. ACC to thalamus effective connectivity had a positive association in chronic pain patients who had experienced emotional neglect in their childhood and a negative association in chronic pain patients who had experienced emotional abuse, see Figure 2(b) and Supplementary Materials. It is noteworthy that there was no significant relationship found between effective connectivity and chronic pain comorbidity, with the severity of depression or scores of ACEs, including sexual abuse, physical abuse, and emotional abuse.

**Figure 2:**
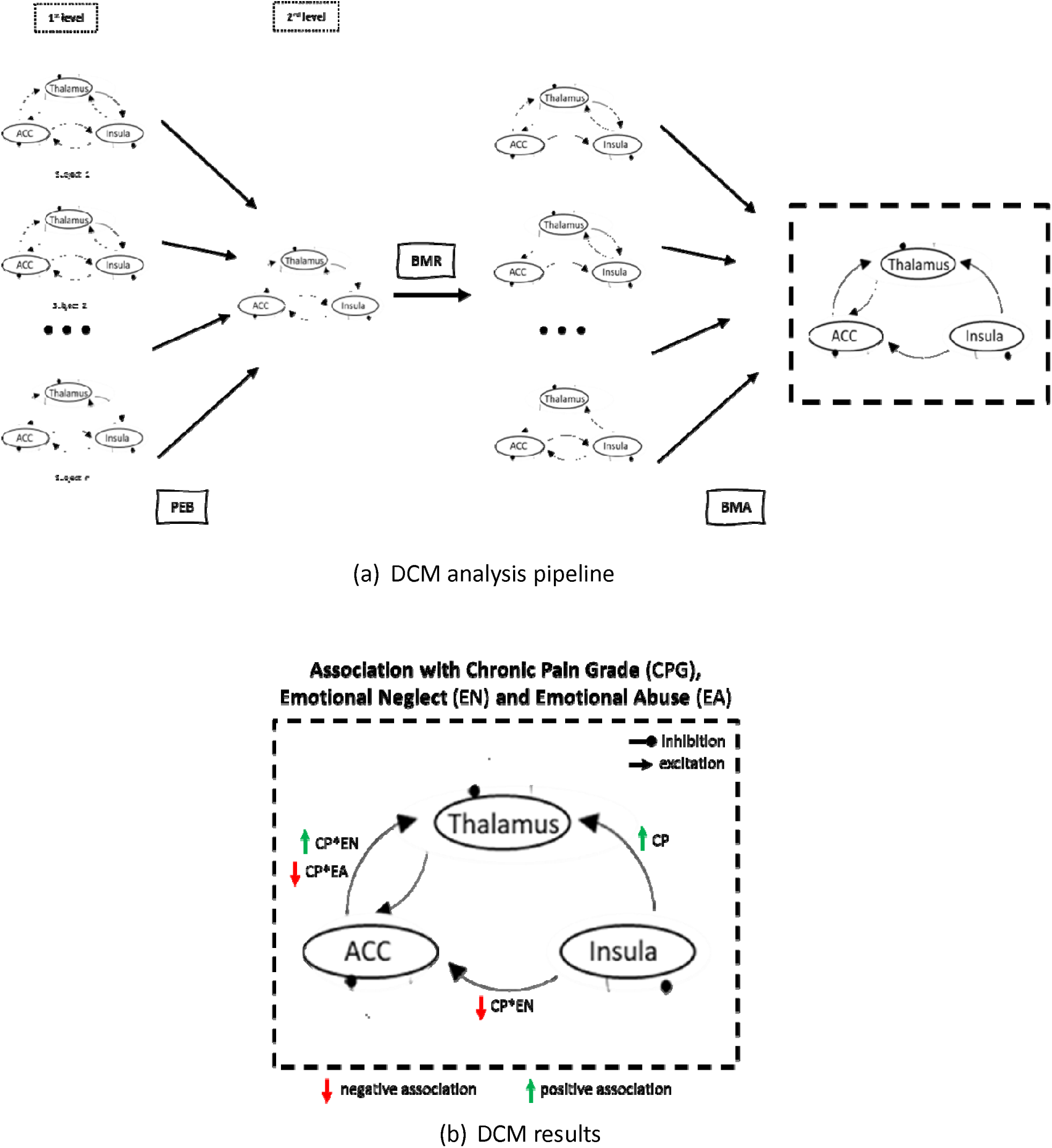
(a) DCMs were brought to the second stage, where the Bayesian Model Reduction (BMR) technique was applied to remove any unnecessary connections. Then, the Bayesian Model Averaging (BMA) method was utilized to combine the DCMs into a single model, taking into account the probabilities of each individual DCM. The final result was a weighted average of the DCMs. (b) Insula to thalamus connection was increased in chronic pain patients. Insula to thalamus connection was decreased in chronic pain (CP) patients with experiences of emotional neglect (EN) in their childhood. ACC to thalamus connection increased in chronic pain patients with experiences of emotional neglect in their childhood and decreased in chronic pain patients with experiences of emotional abuse (EA) in their childhood.

**Figure 3:**
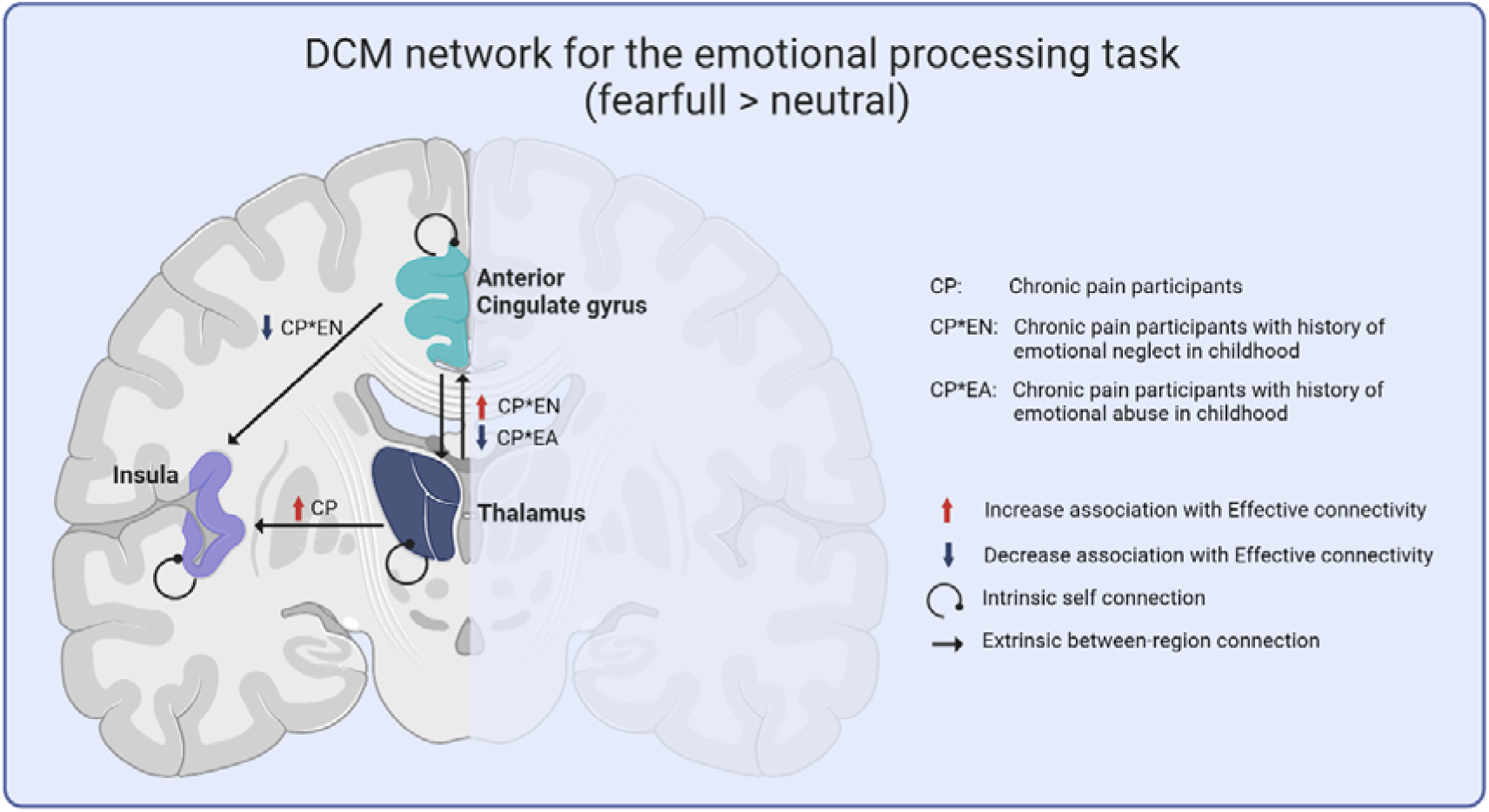
Insula to thalamus connection was increased in chronic pain patients. Insula to thalamus connection was decreased in chronic pain (CP) patients with experiences of emotional neglect (EN) in their childhood. ACC to thalamus connection increased in chronic pain patients with experiences of emotional neglect in their childhood and decreased in chronic pain patients with experiences of emotional abuse (EA) in their childhood. Created with BioRender.com

Further control analyses were conducted to ensure that the results were not influenced by the specific design of the model and the criteria used to determine the variance threshold. In each control analysis, a positive correlation was observed between chronic pain and the effective connectivity from the insula to the thalamus. Regarding the relationship between effective connectivity from the insula to the ACC and chronic pain patients who experienced emotional neglect in childhood, a trend was present, but strong evidence was not consistently found. A similar pattern was found in the relationship between effective connectivity strength from ACC to the thalamus for chronic pain patients who either experienced emotional neglect or emotional abuse in their childhood (Supplementary Materials).

## Discussion

This is the first large, community-based study to demonstrate that histories of adverse events in childhood are associated with altered effective connectivity within the insulo-thalamo-cingulate network in adults with chronic pain. Chronic pain was linked to increased insula → thalamus connectivity, consistent with heightened bottom-up signalling, whereas emotional neglect in childhood was associated with reduced insula → ACC connectivity, indicating weakened top-down regulatory control. ACC → thalamus connectivity differed by ACE subtype, suggesting that emotional neglect and abuse have distinct influences on cortical regulation of thalamic relay. No associations were observed between effective connectivity and depression severity or ACE subtypes, including sexual abuse, physical abuse, and physical neglect, underscoring the specificity of the observed network alterations to certain forms of emotional adversity. Overall, these results indicate that chronic pain and ACEs converge to recalibrate salience network dynamics.

### Bottom-up signalling in chronic pain

Increased insula → thalamus connectivity aligns with models positioning the insula as a hub for integrating interoceptive and nociceptive information into subjective pain perception [7,9,11,29,80]. Heightened bottom-up connectivity may reflect amplified nociceptive processing and salience attribution, consistent with prior findings of hyperactivity in insula and thalamus in chronic pain populations[29,39,66,68,76,80]. Together with the ACC, the anterior insula mediates the emotional and interoceptive aspects of pain[39,68] and coordinates responses to aversive experiences, including social and moral distress[29,66,76]. Demonstrating this in a large cohort strengthens evidence for network-level adaptations in chronic pain.

### Top-down modulation and emotional neglect

Reduced insula → ACC connectivity in individuals with histories of emotional neglect and chronic pain suggests impaired top-down control over salience and affective processing. The ACC is implicated in integrating emotional and cognitive information[13,73,75] and modulating affective responses to nociceptive stimuli[1,77]. Subregions within the ACC have distinct roles: the pregenual ACC modulates negative affect and autonomic responses, whereas the anterior mid-cingulate (posterior ACC) is involved in fear, threat anticipation, and avoidance behaviours[86]. Exposure to ACEs has been linked to structural and functional alterations in the ACC, including reduced grey matter[17] and blunted responses to threat cues[42,89]. Our findings suggest that weakened insula → ACC coupling may limit the capacity to regulate incoming pain signals, providing a mechanistic link between emotional neglect and chronic pain vulnerability.

### Differential effects of adversity types

The ACC → thalamus connectivity patterns varied by chronic pain and type of adversity, with emotional neglect associated with increased coupling and emotional abuse with decreased coupling. This suggests that different forms of early-life adversity differentially influence cortical control over thalamic relay of nociceptive and affective information. Central sensitisation, a heightened state of neural excitability in the central nervous system, which can lead to an increased sensitivity to pain and amplification of pain signals, plays a key role in the development of chronic pain[6]. The thalamus has been shown to become more excitable in patients with chronic pain, which was linked to central sensitisation[61,84] and decreased thalamic grey matter density[4]. Prior work suggests thalamic excitability may be modulated by early adversity[63]. This increased sensitivity could be a result of changes in the thalamus in response to exposure to traumatic experiences, which could lead to the development of chronic pain in adulthood.

Emotional abuse may attenuate top-down regulatory influence, amplifying negative affect and pain, whereas emotional neglect may trigger compensatory strengthening of ACC → thalamus connectivity, reflecting adaptive or maladaptive reorganisation. Such differentiation underscores the importance of considering adversity subtypes when examining neural risk mechanisms for chronic pain. These findings reveal a coherent reorganisation of salience network dynamics in which chronic pain and early-life adversity interact to shape neural processing of sensory and affective signals.

### Strengths and Limitations

ACEs were assessed retrospectively in the current study, and it has been demonstrated that there is only a modest level of agreement between retrospective and prospective measures[72] so recall bias[71] cannot be excluded. Future studies should integrate prospective measures of adversity, such as the Generation Scotland (STRADL-Pain)[36,50,78] and the Avon Longitudinal Study of Parents and Children (ALSPAC)[24]. Additionally, we have not taken into consideration the timing or the length of time that the adversity lasted, even though it could be significantly important[37,74]. Also, the concept of adversity that we and others use is entirely social, meaning it excludes other types of early adversity. Moreover, at the time the imaging took place, there were no available measures of chronic pain in patients.

Several methodological strengths balance these limitations. The large, community-based cohort enhances generalisability and enables robust modelling of ACE- and pain-related network effects. Dynamic causal modelling provides directional, mechanistic insight beyond conventional functional connectivity, while examining multiple ACE subtypes allows identification of adversity-specific network alterations. Finally, ongoing involvement of patient partners throughout the study ensured that the hypotheses and interpretation remained conceptually relevant.

## Conclusion

In summary, chronic pain is associated with increased bottom-up insula → thalamus signalling, while emotional neglect reduces top-down insula → ACC connectivity. ACC → thalamus coupling differs by ACE subtype, highlighting adversity-specific network signatures. These patterns suggest that early-life experiences and chronic pain converge to recalibrate the balance between sensory relay, integrative processing, and cortical regulation, offering a unifying framework for understanding how early experiences embed vulnerability to persistent pain.

## Data Availability

Data are available upon request from the Generation Scotland.

## Details of authors’ contributions

Drafted the manuscript: G.A. Critically revised the manuscript repetitively: G.A., B.S., T.H., L.C. and D.S. Approved the final version of the manuscript: all authors

## Acknowledgements

Generation Scotland received core support from the Chief Scientist Office of the Scottish Government Health Directorates [CZD/16/6] and the Scottish Funding Council [HR03006] and is currently supported by the Wellcome Trust [216767/Z/19/Z].

