## Supplementary material for "From Adversity to Pain: Disrupted Insulo-Cingulo-Thalamic Dynamics in Emotional Abuse and Neglect": suppl

### Dynamic causal modelling of event-related effective connectivity

The mean CPG score was 1.60 for all the participants and the excluded group had mean CPG = 1.61 for the subgroup of participants (Welch's t-test: p = 0.899). The mean Total EA was 5.94 for all the participants and the excluded group had mean Total EA = 6.00 for the subgroup of participants (Welch's t-test: p = 0.640). The mean Total EN was 8.16 for all the participants and the excluded group had mean Total EN = 8.43 for the subgroup of participants (Welch's t-test: p = 0.329). The mean QIDS was 4.58 for all the participants and the excluded group had mean QIDS = 4.61 for the subgroup of participants (Welch's t-test: p = 0.919).

### General linear model voxel-based functional MRI analyses

**De-activation**

| k | T | x | y | z | Region |
|---|---|---|---|---|---|
| 13597 | 9.60 | -8 | 46 | 14 | *L & R ACC* |
| | 9.38 | 8 | 38 | 24 | *L & R Frontal Superior Medial* |
| | 9.34 | 40 | 26 | 42 | *L Middle Cingulate* |
| 24162 | 9.10 | -40 | -64 | -44 | *L Cerebellum* |
| | 9.05 | -4 | -60 | 62 | *L Precuneus* |
| | 8.66 | -34 | -66 | -30 | |
| 141 | 7.15 | 38 | -80 | -26 | *R Cerebellum* |
| | 6.03 | 50 | -62 | -28 | |
| 76 | 7.07 | 28 | -34 | -28 | *R Cerebellum* |
| 427 | 7.00 | -30 | 16 | 6 | *L Insula* |
| | 7.00 | -34 | 10 | -4 | *L Putamen, Caudate* |
| | 6.27 | -24 | 26 | -4 | |
| 143 | 6.97 | -54 | -54 | -14 | *L Inferior Temporal* |
| 126 | 6.46 | -38 | -80 | 26 | *L Middle Occipital* |
| 48 | 6.39 | 30 | -42 | -40 | *R Cerebellum* |
| | 6.13 | 24 | -44 | -50 | |
| 129 | 6.18 | 38 | 16 | 0 | *R Insula* |
| | 5.99 | 32 | 20 | -8 | |
| 70 | 5.96 | -30 | -6 | 48 | *L Precentral* |
| 5 | 5.83 | -8 | -2 | 74 | *L Supplementary Motor Area* |
| 11 | 5.65 | 2 | -18 | 16 | *R Thalamus* |
| 6 | 5.56 | 2 | -22 | -22 | *VTA* |
| 1 | 5.51 | -24 | -66 | 34 | *L Occipital* |
| 2 | 5.49 | 22 | 24 | 12 | *R Caudate, Putamen, Insula* |
| 1 | 5.46 | -24 | -64 | 40 | *L Superior Parietal* |
| 1 | 5.44 | 12 | 38 | 2 | *R ACC* |

*Table 1 Results of significant de-activations regarding fearful and neutral faces across 579 participants. ACC, Anterior Cingulate Cortex; VTA*

**CPG-positive association**

| k | T | x | y | z | Region |
|---|---|---|---|---|---|
| 1597 | 3.24 | 16 | -22 | 50 | *R Middle Cingulate* |
| | 3.11 | 10 | 4 | 18 | *R Supplementary Motor Area* |
| | 2.81 | 28 | -18 | 52 | |
| 1297 | 3.18 | -50 | -30 | 24 | *L Supramarginal* |
| | 2.61 | -12 | -30 | 50 | |
| | 2.61 | -30 | -26 | 38 | |
| 377 | 2.81 | 34 | 10 | 34 | *R Frontal Inferior Operculum* |
| | 2.19 | 20 | 12 | 48 | *R Superior Frontal* |
| | 2.11 | 32 | 0 | 38 | |
| 183 | 2.53 | -36 | 20 | 42 | *L Middle Frontal* |
| | 2.26 | -30 | 20 | 32 | |
| | 2.00 | -36 | 32 | 26 | |
| 396 | 2.51 | -28 | 4 | -8 | *L Putamen* |
| | 2.30 | -32 | -6 | 0 | |
| | 2.15 | -28 | -12 | -6 | |

**CPG-negative association**

| k | T | x | y | z | Region |
|---|---|---|---|---|---|
| 3177 | 3.31 | 66 | -50 | 14 | *R Middle & Superior Temporal* |
| | 3.14 | 38 | -24 | 12 | *R Heschl* |
| | 2.92 | 54 | -34 | 12 | |
| 162 | 3.27 | -6 | 12 | -12 | *Nucleus Accumbens* |
| | 2.08 | -2 | 20 | 6 | |
| 304 | 3.09 | 22 | 26 | 34 | *R Superior Frontal* |
| | 2.43 | 22 | 28 | -2 | |
| | 2.28 | 18 | 30 | 8 | |
| 347 | 3.02 | 0 | -82 | -10 | *L Calcarine* |
| | 1.97 | -10 | -62 | -24 | *L Cerebellum* |
| | 1.84 | -12 | -78 | -10 | |
| 1012 | 3.00 | -48 | -46 | -6 | *L Middle Temporal* |
| | 2.94 | -38 | -62 | 12 | |
| | 2.74 | -58 | -66 | 16 | |
| 231 | 2.84 | -16 | 20 | 66 | *L Superior Frontal* |
| | 2.62 | -30 | 20 | 58 | |
| 160 | 2.59 | -20 | -40 | -18 | *L Cerebellum* |
| | 2.12 | -22 | -28 | -22 | *L Parahippocampal* |
| | 2.03 | -14 | -24 | -28 | |
| 155 | 2.43 | -32 | -64 | 58 | *L Superior Parietal* |
| | 2.10 | -14 | -68 | 64 | |
| | 1.76 | -14 | -60 | 70 | |
| 171 | 2.23 | -24 | -60 | -28 | *L Cerebellum* |
| | 1.99 | -24 | -68 | -22 | |
| 130 | 2.17 | 20 | 48 | 8 | *R Middle Frontal* |
| | 2.16 | 28 | 38 | 6 | |
| | 2.09 | 32 | 54 | 4 | |

*Table 2 Results of significant de-activations regarding fearful and neutral faces and Chronic Pain Grade (CPG) associations. ACC, Anterior Cingulate Cortex; VTA*

**Chronic Pain and SA-positive association**

| k | T | x | y | z | Region |
|---|---|---|---|---|---|
| 4903 | 3.59 | -10 | 4 | 52 | *L Supplementary Motor Area* |
| | 3.44 | -34 | -8 | 22 | |
| | 2.92 | -18 | 12 | 34 | |
| 540 | 3.46 | 24 | 42 | 6 | *R Middle & Superior Frontal* |
| | 2.22 | 20 | 48 | 22 | |
| | 2.19 | 14 | 44 | 28 | |
| 2086 | 3.19 | 60 | 2 | 16 | *R Rolandic Operculum* |
| | 2.81 | 66 | -6 | 18 | *R Postcentral* |
| | 2.79 | 56 | 2 | 24 | |
| 144 | 2.77 | 12 | 44 | 54 | *R Superior Medial frontal* |
| | 2.04 | 18 | 32 | 60 | |
| 269 | 2.70 | -24 | -36 | -36 | *Cerebellum* |
| | 2.56 | -20 | -48 | -28 | |
| 1145 | 2.66 | 20 | -76 | 22 | *R Occipital* |
| | 2.43 | 20 | -62 | 36 | |
| | 2.40 | 26 | -66 | 28 | |
| 1446 | 2.55 | -22 | -64 | 4 | *L Calcarine, Lingual* |
| | 2.47 | -22 | -54 | 2 | |
| | 2.41 | -18 | -82 | 22 | |
| 143 | 2.49 | -24 | 2 | -20 | *L Amygdala* |
| 242 | 2.43 | -24 | 42 | 12 | *L Middle & Superior Frontal* |
| | 2.28 | -28 | 46 | 18 | |
| | 2.24 | -26 | 50 | 34 | |
| 303 | 2.32 | 28 | 2 | 40 | *L Middle & Superior Frontal* |
| | 2.27 | 46 | -10 | 52 | |
| | 1.92 | 36 | -4 | 54 | |

**Chronic Pain and SA-negative association**

| k | T | x | y | z | Region |
|---|---|---|---|---|---|
| - | - | - | - | - | - |

*Table 3 Results of significant de-activations regarding fearful and neutral faces and Chronic Pain with history of sexual abuse (SA) in childhood.*

**Chronic Pain and EA-positive association**

| k | T | x | y | z | Region |
|---|---|---|---|---|---|
| 137 | 3.07 | 38 | -14 | -12 | *R Hippocampus/Amygdala* |
| | 2.12 | 36 | -20 | -4 | |
| 1889 | 2.98 | 14 | -100 | 8 | *Occipital* |
| | 2.84 | 26 | -86 | 16 | |
| | 2.60 | 40 | -64 | 28 | |
| 439 | 2.83 | 56 | -28 | -4 | *R Middle & Superior Temporal* |
| | 2.67 | 54 | -20 | 10 | *R Heschl* |
| | 2.19 | 44 | -22 | 10 | |
| 558 | 2.75 | -40 | -72 | -10 | *Occipital* |
| | 2.53 | -44 | -84 | -8 | |
| | 2.33 | -34 | -92 | -2 | |
| 1705 | 2.71 | 28 | 32 | 28 | *R Middle Frontal* |
| | 2.68 | 52 | -2 | 44 | |
| | 2.67 | 38 | 16 | 30 | |
| 253 | 2.63 | 44 | 14 | -34 | *R Middle Temporal* |
| | 2.48 | 52 | -12 | -22 | |
| | 2.40 | 50 | 2 | -22 | |
| 138 | 2.36 | -10 | 50 | 8 | *L Superior Medial frontal* |
| | 2.14 | -6 | 58 | 16 | |
| | 1.85 | -2 | 64 | 20 | |
| 224 | 2.16 | -52 | -34 | 10 | *L Superior Temporal* |
| | 2.01 | -60 | -26 | 10 | |
| | 2.01 | -52 | -24 | 12 | |

**Chronic Pain and EA-negative association**

| k | T | x | y | z | Region |
|---|---|---|---|---|---|
| 251 | 2.59 | 12 | -24 | 24 | *R middle cingulate* |
| | 2.09 | 14 | -34 | 24 | *R Thalamus* |
| 270 | 2.36 | -14 | -32 | 22 | *L Thalamus* |
| | 2.19 | -24 | -28 | 34 | *PCC* |
| | 1.84 | -6 | -44 | 16 | |

*Table 4 Results of significant de-activations regarding fearful and neutral faces and Chronic Pain with history of emotional abuse (EA) in childhood.*

**Chronic Pain and EN-positive association**

| k | T | x | y | z | Region |
|---|---|---|---|---|---|
| 143 | 2.49 | -14 | 18 | 16 | *L Caudate* |
| 183 | 2.48 | -18 | -10 | -26 | *L Parahippocampal/ Hippocampus* |
| | 2.42 | -24 | 2 | -20 | *L Amygdala* |
| 456 | 2.37 | 24 | -82 | 20 | *Occipital* |
| | 2.22 | 32 | -74 | 18 | |
| | 2.18 | 38 | -68 | 26 | |

**Chronic Pain and EN-negative association**

| k | T | x | y | z | Region |
|---|---|---|---|---|---|
| 840 | 3.27 | -36 | 24 | 38 | *L Middle & Superior Frontal* |
| | 2.67 | -16 | 16 | 66 | *L Supplementary Motor Area* |
| | 2.40 | -26 | 36 | 42 | |
| 1131 | 3.07 | -26 | -62 | -44 | *Cerebellum* |
| | 2.93 | -4 | -46 | -50 | |
| | 2.61 | 6 | -48 | -60 | |
| 987 | 2.99 | 24 | -38 | 10 | *R Hippocampus* |
| | 2.73 | 12 | -26 | 20 | *R Thalamus* |
| | 2.60 | 30 | -32 | 8 | |
| 363 | 2.79 | 38 | 8 | 8 | *R Insula* |
| | 2.45 | 46 | 18 | 8 | *R Putamen* |
| | 2.21 | 54 | 14 | 8 | |
| 942 | 2.70 | -48 | -58 | 12 | *L Middle & Superior Temporal* |
| | 2.43 | -56 | -46 | 16 | |
| | 2.36 | -44 | -46 | 20 | |
| 167 | 2.54 | 48 | 34 | -2 | *R Inferior Frontal* |
| | 2.34 | 40 | 42 | 0 | |
| | 1.78 | 40 | 30 | -6 | |
| 319 | 2.37 | -36 | 18 | -28 | *L Temporal Pole* |
| | 2.37 | -42 | 46 | -2 | |
| | 2.18 | -34 | 48 | -4 | |
| 156 | 2.28 | 12 | 38 | 20 | *R ACC* |
| | 2.13 | 20 | 54 | 14 | |
| | 2.00 | 18 | 62 | 8 | |

*Table 5 Results of significant de-activations regarding fearful and neutral faces and Chronic Pain with history of emotional neglect (EN) in childhood.*

**Chronic Pain and PN-positive association**

| k | T | x | y | z | Region |
|---|---|---|---|---|---|
| 3990 | 3.56 | 28 | 0 | 40 | *R Middle Frontal* |
| | 3.18 | 58 | 0 | 18 | |
| | 3.13 | 58 | 8 | 30 | |
| 162 | 3.02 | -24 | -38 | -38 | *L Cerebellum* |
| | 2.19 | -20 | -46 | -32 | |
| 931 | 2.86 | -54 | -6 | 16 | *L Postcentral* |
| | 2.80 | -52 | -22 | 30 | |
| | 2.69 | -48 | 2 | 8 | |
| 2394 | 2.81 | 28 | -80 | 16 | *R Occipital* |
| | 2.64 | 22 | -68 | 38 | |
| | 2.62 | 24 | -88 | 18 | |
| 441 | 2.66 | -10 | -22 | 36 | *L & R Middle Cingulate* |
| | 2.49 | 4 | -14 | 32 | |
| 150 | 2.62 | 14 | 42 | 30 | *R Superior Frontal* |
| | 1.99 | 26 | 36 | 26 | |
| | 1.90 | 22 | 42 | 32 | |
| 173 | 2.55 | -50 | 12 | -24 | *L Superior temporal* |
| | 2.46 | -40 | 10 | -32 | |
| | 1.78 | -44 | 4 | -38 | |
| 198 | 2.43 | -26 | 4 | -14 | *L Amygdala* |
| | 1.81 | -34 | 10 | -8 | |
| 197 | 2.33 | 4 | -24 | 16 | *R Thalamus* |
| | 2.26 | -6 | -20 | 10 | |
| 139 | 2.21 | -4 | -48 | -2 | *L Cerebellum* |
| | 1.95 | 4 | -48 | -6 | |
| | 1.88 | 8 | -46 | -14 | |
| 176 | 2.21 | -28 | -98 | -4 | *L Occipital* |
| | 2.05 | -24 | -90 | 2 | |
| | 1.83 | -18 | -102 | -2 | |

**Chronic Pain and PN-negative association**

| k | T | x | y | z | Region |
|---|---|---|---|---|---|
| 559 | 3.44 | -46 | -80 | 28 | *L Occipital* |
| | 3.04 | -50 | -74 | 22 | |
| | 2.44 | -48 | -56 | 16 | |
| 161 | 3.10 | -12 | 64 | 6 | *L Superior Medial Frontal* |
| | 2.16 | 6 | 60 | 6 | |
| 253 | 2.80 | 64 | -14 | -8 | *R Superior temporal* |
| | 2.51 | 70 | -20 | -8 | |
| 186 | 2.79 | -24 | -66 | -40 | *L Cerebellum* |
| 190 | 2.35 | 14 | -26 | 20 | *R Thalamus* |
| | 2.22 | 26 | -40 | 14 | |

*Table 6 Results of significant de-activations regarding fearful and neutral faces and Chronic Pain with history of physical neglect (PN) in childhood.*

**Chronic Pain and PA-positive association**

| k | T | x | y | z | Region |
|---|---|---|---|---|---|
| 575 | 3.28 | 12 | -28 | 20 | *R Thalamus* |
| | 1.97 | 16 | -20 | 32 | *R middle cingulate* |
| 226 | 3.06 | -30 | 42 | -2 | *L Middle & Superior Frontal* |
| | 1.81 | -42 | 44 | 0 | |
| 167 | 3.01 | -10 | -36 | -20 | *L Cerebellum* |
| 182 | 2.73 | 0 | -46 | -50 | *R Cerebellum* |
| | 2.60 | -6 | -38 | -56 | |
| | 1.92 | 6 | -48 | -60 | |
| 297 | 2.68 | -16 | -36 | 20 | *L posterior cingulate* |

**Chronic Pain and PA-negative association**

| k | T | x | y | z | Region |
|---|---|---|---|---|---|
| 927 | 3.23 | 54 | -6 | -2 | *R Superior temporal* |
| | 2.84 | 38 | -10 | -12 | *R Hippocampus* |
| | 2.83 | 16 | 2 | 18 | *R Caudate* |
| 2225 | 3.21 | -10 | 2 | 54 | *L Supplementary Motor Area* |
| | 3.13 | 28 | 32 | 28 | *R Middle Frontal* |
| | 2.86 | 26 | 4 | 42 | |
| 2188 | 3.11 | 22 | -62 | 38 | *Occipital* |
| | 3.03 | 24 | -68 | 32 | |
| | 2.79 | 30 | -72 | 20 | |
| 968 | 3.03 | -10 | 14 | 14 | *L Caudate* |
| | 2.63 | -18 | -2 | 18 | *L ACC* |
| | 2.33 | -8 | 20 | 24 | |
| 298 | 2.77 | -18 | -78 | 48 | *L Superior Parietal* |
| | 2.14 | -20 | -68 | 36 | |
| 925 | 2.70 | 52 | -2 | 44 | *R Precentral* |
| | 2.60 | 56 | 4 | 32 | |
| | 2.48 | 32 | -16 | 62 | |
| 334 | 2.60 | -66 | -46 | 16 | *L Superior & Middle Temporal* |
| | 2.43 | -60 | -26 | 10 | |
| | 2.13 | -50 | -34 | 12 | |
| 235 | 2.58 | 2 | -70 | 2 | *R Lingual* |
| | 1.91 | -10 | -66 | 6 | |
| 338 | 2.46 | 4 | -46 | -20 | *Vermis* |
| | 2.17 | -8 | -54 | -18 | |
| | 2.09 | -18 | -50 | -30 | |

*Table 7 Results of significant de-activations regarding fearful and neutral faces and Chronic Pain with history of physical abuse (PA) in childhood.*

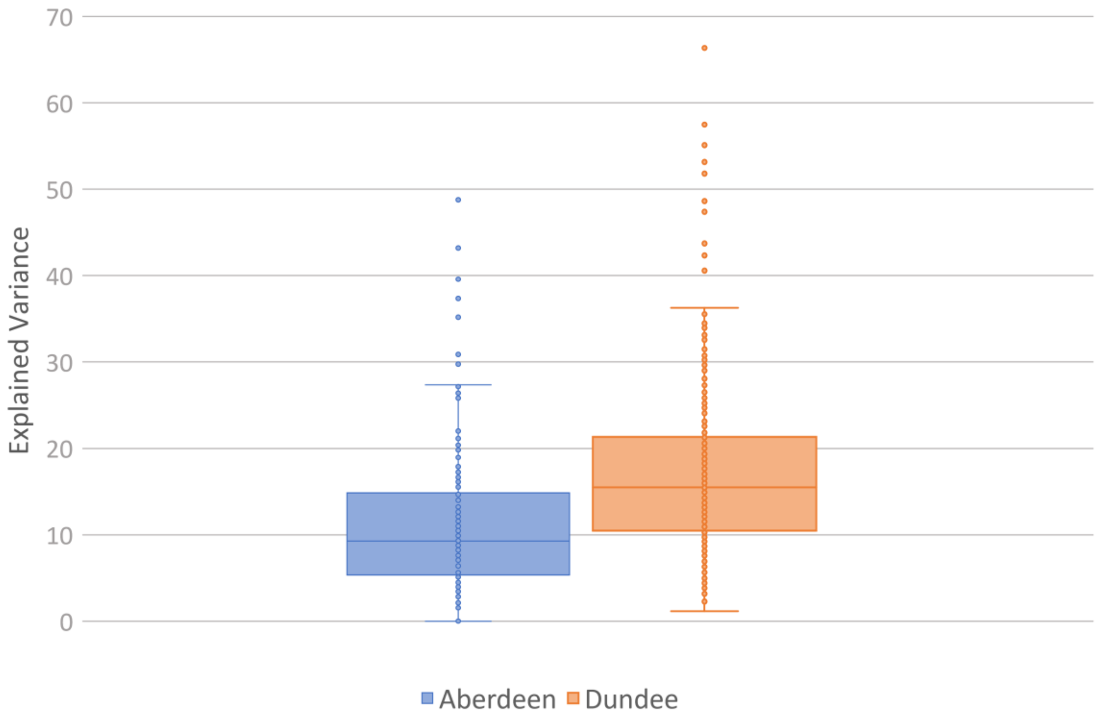


*Suppl. Figure 1 Left Hemisphere – Explained Variance for different sites (Aberdeen and Dundee)*

| PEB: A and B matrix | | All participants - 579 | | | | >=7.5% participants - 457 | | | | >=10% participants - 371 | | | | >=15% participants - 235 | | | |
|---|---|---|---|---|---|---|---|---|---|---|---|---|---|---|---|---|---|
| | | P(common) | P(CP) | E(common) | E(CP) | P(common) | P(CP) | E(common) | E(CP) | P(common) | P(CP) | E(common) | E(CP) | P(common) | P(CP) | E(common) | E(CP) |
| IN -> IN | A (1,1) | 1.00 | 0.00 | -0.34 | 0.00 | 1.00 | 0.00 | -0.40 | 0.00 | 1.00 | 0.00 | -0.42 | 0.00 | 1.00 | 0.00 | -0.44 | 0.00 |
| IN -> ACC | A (2,1) | 1.00 | 0.00 | 0.36 | 0.00 | 1.00 | 0.00 | 0.39 | 0.00 | 1.00 | 0.00 | 0.41 | 0.00 | 1.00 | 0.00 | 0.44 | 0.00 |
| IN -> TH | A (3,1) | 1.00 | 0.83 | 0.25 | 0.09 | 1.00 | 0.97 | 0.24 | 0.14 | 1.00 | 0.87 | 0.25 | 0.11 | 1.00 | 0.63 | 0.24 | 0.07 |
| ACC -> IN | A (1,2) | 0.00 | 0.00 | 0.00 | 0.00 | 0.00 | 0.00 | 0.00 | 0.00 | 0.00 | 0.00 | 0.00 | 0.00 | 0.00 | 0.00 | 0.00 | 0.00 |
| ACC -> ACC | A (2,2) | 1.00 | 0.51 | -0.46 | 0.04 | 1.00 | 0.00 | -0.51 | 0.00 | 1.00 | 0.63 | -0.54 | 0.05 | 1.00 | 0.00 | -0.60 | 0.00 |
| ACC -> TH | A (3,2) | 1.00 | 0.00 | 0.11 | 0.00 | 1.00 | 0.00 | 0.11 | 0.00 | 1.00 | 0.00 | 0.11 | 0.00 | 1.00 | 0.00 | 0.12 | 0.00 |
| TH -> IN | A (1,3) | 0.00 | 0.00 | 0.00 | 0.00 | 0.00 | 0.00 | 0.00 | 0.00 | 0.00 | 0.00 | 0.00 | 0.00 | 0.00 | 0.00 | 0.00 | 0.00 |
| TH -> ACC | A (2,3) | 1.00 | 0.00 | -0.05 | 0.00 | 1.00 | 0.00 | -0.08 | 0.00 | 1.00 | 0.00 | -0.08 | 0.00 | 1.00 | 0.00 | -0.11 | 0.00 |
| TH -> TH | A (3,3) | 1.00 | 0.00 | -0.30 | 0.00 | 1.00 | 0.00 | -0.34 | 0.00 | 1.00 | 0.00 | -0.37 | 0.00 | 1.00 | 0.00 | -0.42 | 0.00 |
| IN -> ACC | B (2,1,3) | 1.00 | 0.00 | 0.41 | 0.00 | 1.00 | 0.00 | 0.39 | 0.00 | 1.00 | 0.00 | 0.36 | 0.00 | 0.99 | 0.00 | 0.30 | 0.00 |
| IN -> TH | B (3,1,3) | 1.00 | 0.58 | 0.35 | -0.22 | 0.94 | 0.00 | 0.24 | 0.00 | 0.69 | 0.00 | 0.14 | 0.00 | 1.00 | 0.00 | 0.38 | 0.00 |
| ACC -> IN | B (1,2,3) | 1.00 | 0.00 | 1.65 | 0.00 | 1.00 | 0.00 | 1.74 | 0.00 | 1.00 | 0.00 | 1.72 | 0.00 | 1.00 | 0.00 | 1.65 | 0.00 |
| ACC -> TH | B (3,2,3) | 1.00 | 0.00 | 0.85 | 0.00 | 1.00 | 0.00 | 0.85 | 0.00 | 1.00 | 0.00 | 0.88 | 0.00 | 1.00 | 0.00 | 0.70 | 0.00 |
| TH -> IN | B (1,3,3) | 1.00 | 0.00 | -0.65 | 0.00 | 1.00 | 0.00 | -0.74 | 0.00 | 1.00 | 0.00 | -0.78 | 0.00 | 1.00 | 0.00 | -0.79 | 0.00 |
| TH -> ACC | B (2,3,3) | 0.90 | 0.00 | -0.15 | 0.00 | 0.79 | 0.00 | -0.13 | 0.00 | 0.70 | 0.00 | -0.11 | 0.00 | 0.00 | 0.00 | 0.00 | 0.00 |

*Table 8 **BMA results of intrinsic and extrinsic using a PEB matrix without covariates.** P(x) shows the posterior probability of the estimates (E(x)) being non-zero. The first column identifies the connection as A(TO,FROM), where 1=insula, 2=anterior cingulate cortex, 3=thalamus; e.g. A(1,2) is the connection from ACC to insula.*

| **PEB: A matrix** | | **All** participants - 579 | | | | **>=7.5%** participants - 457 | | | | **>=10%** participants - 371 | | | | **>=15%** participants - 235 | | | |
|---|---|---|---|---|---|---|---|---|---|---|---|---|---|---|---|---|---|
| | | P(common) | P(CP) | E(common) | E(CP) | P(common) | P(CP) | E(common) | E(CP) | P(common) | P(CP) | E(common) | E(CP) | P(common) | P(CP) | E(common) | E(CP) |
| IN -> IN | A (1,1) | 1.00 | 0.53 | -0.38 | -0.04 | 1.00 | 0.00 | -0.43 | 0.00 | 1.00 | 0.00 | -0.44 | 0.00 | 1.00 | 0.00 | -0.46 | 0.00 |
| IN -> ACC | A (2,1) | 1.00 | 0.00 | 0.38 | 0.00 | 1.00 | 0.00 | 0.42 | 0.00 | 1.00 | 0.00 | 0.44 | 0.00 | 1.00 | 0.00 | 0.46 | 0.00 |
| IN -> TH | A (3,1) | 1.00 | 0.90 | 0.27 | 0.10 | 1.00 | 0.98 | 0.27 | 0.14 | 1.00 | 0.90 | 0.27 | 0.12 | 1.00 | 0.75 | 0.25 | 0.10 |
| ACC -> IN | A (1,2) | 0.00 | 0.00 | 0.00 | 0.00 | 0.00 | 0.00 | 0.00 | 0.00 | 0.00 | 0.00 | 0.00 | 0.00 | 0.00 | 0.00 | 0.00 | 0.00 |
| ACC -> ACC | A (2,2) | 1.00 | 0.52 | -0.44 | 0.04 | 1.00 | 0.00 | -0.50 | 0.00 | 1.00 | 0.65 | -0.54 | 0.07 | 1.00 | 0.00 | -0.59 | 0.00 |
| ACC -> TH | A (3,2) | 1.00 | 0.00 | 0.12 | 0.00 | 1.00 | 0.00 | 0.11 | 0.00 | 1.00 | 0.00 | 0.11 | 0.00 | 1.00 | 0.00 | 0.11 | 0.00 |
| TH -> IN | A (1,3) | 0.00 | 0.00 | 0.00 | 0.00 | 0.00 | 0.00 | 0.00 | 0.00 | 0.00 | 0.00 | 0.00 | 0.00 | 0.00 | 0.00 | 0.00 | 0.00 |
| TH -> ACC | A (2,3) | 0.99 | 0.00 | -0.05 | 0.00 | 1.00 | 0.00 | -0.07 | 0.00 | 1.00 | 0.00 | -0.08 | 0.00 | 1.00 | 0.00 | -0.12 | 0.00 |
| TH -> TH | A (3,3) | 1.00 | 0.00 | -0.32 | 0.00 | 1.00 | 0.00 | -0.35 | 0.00 | 1.00 | 0.00 | -0.38 | 0.00 | 1.00 | 0.00 | -0.43 | 0.00 |

*Table 9* ***BMA results of intrinsic connections using a PEB matrix without covariates****. P(x) shows the posterior probability of the estimates (E(x)) being non-zero. The first column identifies the connection as A(TO,FROM), where 1=insula, 2=anterior cingulate cortex, 3=thalamus; e.g. A(1,2) is the connection from ACC to insula.*

| **PEB: B matrix** | | **All** participants - 579 | | | | **>=7.5%** participants - 457 | | | | **>=10%** participants - 371 | | | | **>=15%** participants - 235 | | | |
|---|---|---|---|---|---|---|---|---|---|---|---|---|---|---|---|---|---|
| | | P(common) | P(CP) | E(common) | E(CP) | P(common) | P(CP) | E(common) | E(CP) | P(common) | P(CP) | E(common) | E(CP) | P(common) | P(CP) | E(common) | E(CP) |
| IN -> ACC | B (2,1,3) | 1.00 | 0.00 | 0.73 | 0.00 | 1.00 | 0.00 | 0.73 | 0.00 | 1.00 | 0.00 | 0.66 | 0.00 | 1.00 | 0.00 | 0.57 | 0.00 |
| IN -> TH | B (3,1,3) | 0.00 | 0.00 | 0.00 | 0.00 | 0.00 | 0.00 | 0.00 | 0.00 | 0.00 | 0.00 | 0.00 | 0.00 | 0.00 | 0.00 | 0.00 | 0.00 |
| ACC -> IN | B (1,2,3) | 1.00 | 0.00 | 1.82 | 0.00 | 1.00 | 0.00 | 1.91 | 0.00 | 1.00 | 0.00 | 1.89 | 0.00 | 1.00 | 0.00 | 1.88 | 0.00 |
| ACC -> TH | B (3,2,3) | 1.00 | 0.00 | 1.28 | 0.00 | 1.00 | 0.00 | 1.26 | 0.00 | 1.00 | 0.00 | 1.23 | 0.00 | 1.00 | 0.00 | 1.02 | 0.00 |
| TH -> IN | B (1,3,3) | 1.00 | 0.00 | -0.68 | 0.00 | 1.00 | 0.00 | -0.78 | 0.00 | 1.00 | 0.00 | -0.86 | 0.00 | 1.00 | 0.00 | -0.84 | 0.00 |
| TH -> ACC | B (2,3,3) | 0.00 | 0.00 | 0.00 | 0.00 | 0.00 | 0.00 | 0.00 | 0.00 | 0.00 | 0.00 | 0.00 | 0.00 | 0.00 | 0.00 | 0.00 | 0.00 |

*Table 10* ***BMA results of extrinsic connections using a PEB matrix without covariates****. P(x) shows the posterior probability of the estimates (E(x)) being non-zero. The first column identifies the connection as A(TO,FROM), where 1=insula, 2=anterior cingulate cortex, 3=thalamus; e.g. A(1,2) is the connection from ACC to insula.*

| **PEB: A matrix** | | **No excluded variance threshold** (participants – 579) | | | | | | | | | |
|---|---|---|---|---|---|---|---|---|---|---|---|
| | | P(**common**) | P(**CP**) | P(**CP*EA**) | P(**CP*EN**) | P(**Sex-F1**) | E(**common**) | E(**CP**) | E(**CP*EA**) | E(**CP*EN**) | E(**Sex-F1**) |
| IN -> IN | A (1,1) | 1.00 | 0.53 | 0.00 | 0.00 | 0.00 | -0.38 | -0.04 | 0.00 | 0.00 | 0.00 |
| IN -> ACC | A (2,1) | 1.00 | 0.00 | 0.00 | 0.63 | 0.88 | 0.38 | 0.00 | 0.00 | -0.09 | 0.08 |
| IN -> TH | A (3,1) | 1.00 | 0.90 | 0.00 | 0.00 | 0.00 | 0.27 | 0.10 | 0.00 | 0.00 | 0.00 |
| ACC -> IN | A (1,2) | 0.00 | 0.00 | 0.00 | 0.00 | 0.00 | 0.00 | 0.00 | 0.00 | 0.00 | 0.00 |
| ACC -> ACC | A (2,2) | 1.00 | 0.54 | 0.00 | 0.00 | 0.00 | -0.44 | 0.05 | 0.00 | 0.00 | 0.00 |
| ACC -> TH | A (3,2) | 1.00 | 0.00 | 0.00 | 0.00 | 0.00 | 0.12 | 0.00 | 0.00 | 0.00 | 0.00 |
| TH -> IN | A (1,3) | 0.00 | 0.00 | 0.00 | 0.00 | 0.00 | 0.00 | 0.00 | 0.00 | 0.00 | 0.00 |
| TH -> ACC | A (2,3) | 0.99 | 0.00 | 0.42 | 0.64 | 0.00 | -0.05 | 0.00 | -0.05 | 0.09 | 0.00 |
| TH -> TH | A (3,3) | 1.00 | 0.00 | 0.00 | 0.00 | 0.00 | -0.32 | 0.00 | 0.00 | 0.00 | 0.00 |
| **PEB: A matrix** | | **>=7.5% variance threshold** (participants – 457) | | | | | | | | | |
| | | P(**common**) | P(**CP**) | P(**CP*EA**) | P(**CP*EN**) | P(**Sex-F1**) | E(**common**) | E(**CP**) | E(**CP*EA**) | E(**CP*EN**) | E(**Sex-F1**) |
| IN -> IN | A (1,1) | 1.00 | 0.00 | 0.00 | 0.00 | 0.00 | -0.43 | 0.00 | 0.00 | 0.00 | 0.00 |
| IN -> ACC | A (2,1) | 1.00 | 0.00 | 0.00 | 0.69 | 0.84 | 0.42 | 0.00 | 0.00 | -0.12 | 0.08 |
| IN -> TH | A (3,1) | 1.00 | 0.98 | 0.00 | 0.00 | 0.00 | 0.27 | 0.14 | 0.00 | 0.00 | 0.00 |
| ACC -> IN | A (1,2) | 0.00 | 0.00 | 0.00 | 0.00 | 0.00 | 0.00 | 0.00 | 0.00 | 0.00 | 0.00 |
| ACC -> ACC | A (2,2) | 1.00 | 0.00 | 0.00 | 0.00 | 0.00 | -0.50 | 0.00 | 0.00 | 0.00 | 0.00 |
| ACC -> TH | A (3,2) | 1.00 | 0.00 | 0.87 | 0.83 | 0.00 | 0.11 | 0.00 | -0.24 | 0.23 | 0.00 |
| TH -> IN | A (1,3) | 0.00 | 0.00 | 0.00 | 0.00 | 0.00 | 0.00 | 0.00 | 0.00 | 0.00 | 0.00 |
| TH -> ACC | A (2,3) | 1.00 | 0.00 | 0.00 | 0.00 | 0.00 | -0.07 | 0.00 | 0.00 | 0.00 | 0.00 |
| TH -> TH | A (3,3) | 1.00 | 0.54 | 0.59 | 0.00 | 0.00 | -0.35 | 0.05 | -0.11 | 0.00 | 0.00 |
| **PEB: A matrix** | | **>=10% variance threshold** (participants – 371) | | | | | | | | | |
| | | P(**common**) | P(**CP**) | P(**CP*EA**) | P(**CP*EN**) | P(**Sex-F1**) | E(**common**) | E(**CP**) | E(**CP*EA**) | E(**CP*EN**) | E(**Sex-F1**) |
| IN -> IN | A (1,1) | 1.00 | 0.00 | 0.00 | 0.00 | 0.00 | -0.44 | 0.00 | 0.00 | 0.00 | 0.00 |
| IN -> ACC | A (2,1) | 1.00 | 0.00 | 0.74 | 1.00 | 0.00 | 0.44 | 0.00 | 0.15 | -0.27 | 0.00 |

| | | | | | | | | | | | |
|---|---|---|---|---|---|---|---|---|---|---|---|
| IN -> TH | A (3,1) | 1.00 | 1.00 | 0.00 | 0.00 | 0.00 | 0.27 | 0.14 | 0.00 | 0.00 | 0.00 |
| ACC -> IN | A (1,2) | 0.00 | 0.00 | 0.00 | 0.00 | 0.00 | 0.00 | 0.00 | 0.00 | 0.00 | 0.00 |
| ACC -> ACC | A (2,2) | 1.00 | 0.74 | 0.71 | 0.53 | 0.00 | -0.54 | 0.09 | -0.18 | 0.12 | 0.00 |
| ACC -> TH | A (3,2) | 1.00 | 0.00 | 1.00 | 1.00 | 0.00 | 0.11 | 0.00 | -0.31 | 0.31 | 0.00 |
| TH -> IN | A (1,3) | 0.00 | 0.00 | 0.00 | 0.00 | 0.00 | 0.00 | 0.00 | 0.00 | 0.00 | 0.00 |
| TH -> ACC | A (2,3) | 1.00 | 0.00 | 0.54 | 0.57 | 0.00 | -0.08 | 0.00 | -0.09 | 0.09 | 0.00 |
| TH -> TH | A (3,3) | 1.00 | 0.49 | 0.67 | 0.00 | 0.00 | -0.38 | 0.04 | -0.15 | 0.00 | 0.00 |

| **PEB: A matrix** | | **>=15% variance threshold** (participants – 235) | | | | | | | | | |
|---|---|---|---|---|---|---|---|---|---|---|---|
| | | P(**common**) | P(**CP**) | P(**CP*EA**) | P(**CP*EN**) | P(**Sex-F1**) | E(**common**) | E(**CP**) | E(**CP*EA**) | E(**CP*EN**) | E(**Sex-F1**) |
| IN -> IN | A (1,1) | 1.00 | 0.00 | 0.00 | 0.00 | 0.00 | -0.46 | 0.00 | 0.00 | 0.00 | 0.00 |
| IN -> ACC | A (2,1) | 1.00 | 0.00 | 0.72 | 1.00 | 0.00 | 0.46 | 0.00 | 0.17 | -0.36 | 0.00 |
| IN -> TH | A (3,1) | 1.00 | 0.76 | 0.00 | 0.00 | 0.00 | 0.25 | 0.10 | 0.00 | 0.00 | 0.00 |
| ACC -> IN | A (1,2) | 0.00 | 0.00 | 0.58 | 0.00 | 0.00 | 0.00 | 0.00 | -0.10 | 0.00 | 0.00 |
| ACC -> ACC | A (2,2) | 1.00 | 0.52 | 1.00 | 1.00 | 0.00 | -0.59 | 0.05 | -0.47 | 0.29 | 0.00 |
| ACC -> TH | A (3,2) | 1.00 | 0.00 | 0.00 | 0.00 | 0.00 | 0.12 | 0.00 | 0.00 | 0.00 | 0.00 |
| TH -> IN | A (1,3) | 0.00 | 0.00 | 0.79 | 0.00 | 0.00 | 0.00 | 0.00 | 0.18 | 0.00 | 0.00 |
| TH -> ACC | A (2,3) | 1.00 | 0.55 | 1.00 | 0.00 | 0.00 | -0.12 | 0.04 | -0.28 | 0.00 | 0.00 |
| TH -> TH | A (3,3) | 1.00 | 0.61 | 0.67 | 0.00 | 0.00 | -0.43 | 0.07 | -0.18 | 0.00 | 0.00 |

*Table 11* ***BMA results of intrinsic connections using a PEB matrix with multiple covariates, CP*EA (Chronic pain and history of emotional abuse), CP*EN (Chronic pain and history of emotional neglect) and SEX-F1 (female coded as 1)****. P(x) shows the posterior probability of the estimates (E(x)) being non-zero. The first column identifies the connection as A(TO,FROM), where 1=insula, 2=anterior cingulate cortex, 3=thalamus; e.g. A(1,2) is the connection from ACC to insula.*

### Dundee only results

| PEB: A and B matrix | | **All** participants - 355 | | | | **>=7.5%** participants - 305 | | | | **>=10%** participants - 266 | | | | **>=15%** participants - 176 | | | |
|---|---|---|---|---|---|---|---|---|---|---|---|---|---|---|---|---|---|
| | | P(common) | P(CP) | E(common) | E(CP) | P(common) | P(CP) | E(common) | E(CP) | P(common) | P(CP) | E(common) | E(CP) | P(common) | P(CP) | E(common) | E(CP) |
| IN -> IN | A (1,1) | 1.00 | 0.00 | -0.56 | 0.00 | 1.00 | 0.00 | -0.58 | 0.00 | 1.00 | 0.00 | -0.61 | 0.00 | 1.00 | 0.00 | -0.68 | 0.00 |
| IN -> ACC | A (2,1) | 1.00 | 0.00 | 0.44 | 0.00 | 1.00 | 0.00 | 0.45 | 0.00 | 1.00 | 0.00 | 0.45 | 0.00 | 1.00 | 0.00 | 0.47 | 0.00 |
| IN -> TH | A (3,1) | 1.00 | 0.95 | 0.24 | 0.12 | 1.00 | 0.99 | 0.23 | 0.16 | 1.00 | 0.98 | 0.24 | 0.16 | 1.00 | 0.66 | 0.22 | 0.08 |
| ACC -> IN | A (1,2) | 0.00 | 0.00 | 0.00 | 0.00 | 0.00 | 0.00 | 0.00 | 0.00 | 0.00 | 0.00 | 0.00 | 0.00 | 0.69 | 0.00 | 0.03 | 0.00 |
| ACC -> ACC | A (2,2) | 1.00 | 0.00 | -0.68 | 0.00 | 1.00 | 0.00 | -0.68 | 0.00 | 1.00 | 0.00 | -0.71 | 0.00 | 1.00 | 0.00 | -0.74 | 0.00 |
| ACC -> TH | A (3,2) | 1.00 | 0.00 | 0.11 | 0.00 | 1.00 | 0.00 | 0.12 | 0.00 | 1.00 | 0.00 | 0.11 | 0.00 | 1.00 | 0.00 | 0.12 | 0.00 |
| TH -> IN | A (1,3) | 0.00 | 0.00 | 0.00 | 0.00 | 0.64 | 0.56 | -0.03 | 0.04 | 0.73 | 0.52 | -0.03 | 0.04 | 0.99 | 0.00 | -0.07 | 0.00 |
| TH -> ACC | A (2,3) | 1.00 | 0.00 | -0.08 | 0.00 | 1.00 | 0.00 | -0.09 | 0.00 | 1.00 | 0.00 | -0.09 | 0.00 | 1.00 | 0.00 | -0.12 | 0.00 |
| TH -> TH | A (3,3) | 1.00 | 0.00 | -0.38 | 0.00 | 1.00 | 0.00 | -0.39 | 0.00 | 1.00 | 0.00 | -0.40 | 0.00 | 1.00 | 0.00 | -0.43 | 0.00 |
| IN -> ACC | B (2,1,3) | 1.00 | 0.00 | 0.43 | 0.00 | 1.00 | 0.00 | 0.45 | 0.00 | 1.00 | 0.00 | 0.48 | 0.00 | 1.00 | 0.00 | 0.48 | 0.00 |
| IN -> TH | B (3,1,3) | 1.00 | 0.00 | 0.32 | 0.00 | 1.00 | 0.00 | 0.33 | 0.00 | 1.00 | 0.00 | 0.32 | 0.00 | 1.00 | 0.00 | 0.46 | 0.00 |
| ACC -> IN | B (1,2,3) | 1.00 | 0.00 | 2.03 | 0.00 | 1.00 | 0.00 | 2.03 | 0.00 | 1.00 | 0.00 | 2.01 | 0.00 | 1.00 | 0.00 | 1.93 | 0.00 |
| ACC -> TH | B (3,2,3) | 1.00 | 0.00 | 0.92 | 0.00 | 1.00 | 0.00 | 0.90 | 0.00 | 1.00 | 0.00 | 0.90 | 0.00 | 1.00 | 0.00 | 0.73 | 0.00 |
| TH -> IN | B (1,3,3) | 1.00 | 0.00 | -1.08 | 0.00 | 1.00 | 0.00 | -1.07 | 0.00 | 1.00 | 0.00 | -1.05 | 0.00 | 1.00 | 0.00 | -1.00 | 0.00 |
| TH -> ACC | B (2,3,3) | 1.00 | 0.00 | -0.31 | 0.00 | 1.00 | 0.00 | -0.30 | 0.00 | 1.00 | 0.00 | -0.27 | 0.00 | 1.00 | 0.00 | -0.30 | 0.00 |

*Table 12* ***BMA results of intrinsic and extrinsic using a PEB matrix without covariates using only data acquired in Dundee.*** *P(x) shows the posterior probability of the estimates (E(x)) being non-zero. The first column identifies the connection as A(TO,FROM), where 1=insula, 2=anterior cingulate cortex, 3=thalamus; e.g. A(1,2) is the connection from ACC to insula.*

| **PEB: A matrix** | | All participants - 355 | | | | >=7.5% participants - 305 | | | | >=10% participants - 266 | | | | >=15% participants - 176 | | | |
|---|---|---|---|---|---|---|---|---|---|---|---|---|---|---|---|---|---|
| | | P(common) | P(CP) | E(common) | E(CP) | P(common) | P(CP) | E(common) | E(CP) | P(common) | P(CP) | E(common) | E(CP) | P(common) | P(CP) | E(common) | E(CP) |
| IN -> IN | A (1,1) | 1.00 | 0.00 | -0.60 | 0.00 | 1.00 | 0.00 | -0.61 | 0.00 | 1.00 | 0.00 | -0.64 | 0.00 | 1.00 | 0.00 | -0.70 | 0.00 |
| IN -> ACC | A (2,1) | 1.00 | 0.00 | 0.48 | 0.00 | 1.00 | 0.00 | 0.48 | 0.00 | 1.00 | 0.00 | 0.48 | 0.00 | 1.00 | 0.00 | 0.50 | 0.00 |
| IN -> TH | A (3,1) | 1.00 | 0.96 | 0.27 | 0.13 | 1.00 | 0.99 | 0.26 | 0.17 | 1.00 | 0.99 | 0.27 | 0.17 | 1.00 | 0.73 | 0.24 | 0.10 |
| ACC -> IN | A (1,2) | 0.00 | 0.00 | 0.00 | 0.00 | 0.00 | 0.00 | 0.00 | 0.00 | 0.00 | 0.00 | 0.00 | 0.00 | 0.73 | 0.00 | 0.03 | 0.00 |
| ACC -> ACC | A (2,2) | 1.00 | 0.00 | -0.66 | 0.00 | 1.00 | 0.00 | -0.67 | 0.00 | 1.00 | 0.00 | -0.70 | 0.00 | 1.00 | 0.00 | -0.74 | 0.00 |
| ACC -> TH | A (3,2) | 1.00 | 0.00 | 0.11 | 0.00 | 1.00 | 0.00 | 0.11 | 0.00 | 1.00 | 0.00 | 0.11 | 0.00 | 1.00 | 0.00 | 0.12 | 0.00 |
| TH -> IN | A (1,3) | 0.00 | 0.00 | 0.00 | 0.00 | 0.00 | 0.00 | 0.00 | 0.00 | 0.50 | 0.00 | -0.02 | 0.00 | 0.95 | 0.00 | -0.06 | 0.00 |
| TH -> ACC | A (2,3) | 1.00 | 0.00 | -0.08 | 0.00 | 1.00 | 0.00 | -0.09 | 0.00 | 1.00 | 0.00 | -0.10 | 0.00 | 1.00 | 0.00 | -0.13 | 0.00 |
| TH -> TH | A (3,3) | 1.00 | 0.00 | -0.38 | 0.00 | 1.00 | 0.00 | -0.39 | 0.00 | 1.00 | 0.00 | -0.40 | 0.00 | 1.00 | 0.00 | -0.42 | 0.00 |

*Table 13* ***BMA results of intrinsic connections using a PEB matrix without covariates using only data acquired in Dundee****. P(x) shows the posterior probability of the estimates (E(x)) being non-zero. The first column identifies the connection as A(TO,FROM), where 1=insula, 2=anterior cingulate cortex, 3=thalamus; e.g. A(1,2) is the connection from ACC to insula.*

| **PEB: B matrix** | | All participants - 355 | | | | >=7.5% participants - 305 | | | | >=10% participants - 266 | | | | >=15% participants - 176 | | | |
|---|---|---|---|---|---|---|---|---|---|---|---|---|---|---|---|---|---|
| | | P(common) | P(CP) | E(common) | E(CP) | P(common) | P(CP) | E(common) | E(CP) | P(common) | P(CP) | E(common) | E(CP) | P(common) | P(CP) | E(common) | E(CP) |
| in -> acc | B (2,1,3) | 1.00 | 0.00 | 0.84 | 0.00 | 1.00 | 0.00 | 0.84 | 0.00 | 1.00 | 0.00 | 0.87 | 0.00 | 1.00 | 0.00 | 0.82 | 0.00 |
| in -> th | B (3,1,3) | 0.00 | 0.93 | 0.00 | -0.48 | 0.00 | 0.97 | 0.00 | -0.54 | 0.00 | 0.98 | 0.00 | -0.60 | 0.60 | 0.00 | 0.12 | 0.00 |
| acc -> in | B (1,2,3) | 1.00 | 0.00 | 2.38 | 0.00 | 1.00 | 0.00 | 2.34 | 0.00 | 1.00 | 0.00 | 2.33 | 0.00 | 1.00 | 0.00 | 2.28 | 0.00 |
| acc -> th | B (3,2,3) | 1.00 | 0.00 | 1.26 | 0.00 | 1.00 | 0.00 | 1.24 | 0.00 | 1.00 | 0.00 | 1.22 | 0.00 | 1.00 | 0.00 | 0.98 | 0.00 |
| th -> in | B (1,3,3) | 1.00 | 0.00 | -1.21 | 0.00 | 1.00 | 0.00 | -1.18 | 0.00 | 1.00 | 0.00 | -1.16 | 0.00 | 1.00 | 0.00 | -1.07 | 0.00 |
| th -> acc | B (2,3,3) | 1.00 | 0.00 | -0.48 | 0.00 | 1.00 | 0.00 | -0.47 | 0.00 | 1.00 | 0.00 | -0.43 | 0.00 | 1.00 | 0.00 | -0.41 | 0.00 |

*Table 14* ***BMA results of extrinsic connections using a PEB matrix without covariates using only data acquired in Dundee****. P(x) shows the posterior probability of the estimates (E(x)) being non-zero. The first column identifies the connection as A(TO,FROM), where 1=insula, 2=anterior cingulate cortex, 3=thalamus; e.g. A(1,2) is the connection from ACC to insula.*

| PEB: A matrix | | ALL participants - 335 | | | | | | | | | |
|---|---|---|---|---|---|---|---|---|---|---|---|
| | | | P(**CP**) | P(**CP*EA**) | P(**CP*EN**) | P(**Sex-F1**) | E(**common**) | E(**CP**) | E(**CP*EA**) | E(**CP*EN**) | E(**Sex-F1**) |
| IN -> IN | A (1,1) | 1.00 | 0.00 | 0.00 | 0.00 | 0.00 | -0.60 | 0.00 | 0.00 | 0.00 | 0.00 |
| IN -> ACC | A (2,1) | 1.00 | 0.00 | 0.00 | 0.97 | 0.69 | 0.47 | 0.00 | 0.00 | -0.19 | 0.05 |
| IN -> TH | A (3,1) | 1.00 | 0.96 | 0.00 | 0.00 | 0.00 | 0.27 | 0.14 | 0.00 | 0.00 | 0.00 |
| ACC -> IN | A (1,2) | 0.00 | 0.00 | 0.00 | 0.00 | 0.00 | 0.00 | 0.00 | 0.00 | 0.00 | 0.00 |
| ACC -> ACC | A (2,2) | 1.00 | 0.00 | 0.00 | 0.00 | 0.64 | -0.66 | 0.00 | 0.00 | 0.00 | 0.05 |
| ACC -> TH | A (3,2) | 1.00 | 0.00 | 0.82 | 0.76 | 0.00 | 0.11 | 0.00 | -0.19 | 0.18 | 0.00 |
| TH -> IN | A (1,3) | 0.00 | 0.00 | 0.52 | 0.00 | 0.00 | 0.00 | 0.00 | 0.06 | 0.00 | 0.00 |
| TH -> ACC | A (2,3) | 1.00 | 0.00 | 0.00 | 0.00 | 0.00 | -0.08 | 0.00 | 0.00 | 0.00 | 0.00 |
| TH -> TH | A (3,3) | 1.00 | 0.00 | 0.00 | 0.00 | 0.00 | -0.38 | 0.00 | 0.00 | 0.00 | 0.00 |
| **PEB: A matrix** | | **>=7.5% participants - 305** | | | | | | | | | |
| | | | P(**CP**) | P(**CP*EA**) | P(**CP*EN**) | P(**Sex-F1**) | E(**common**) | E(**CP**) | E(**CP*EA**) | E(**CP*EN**) | E(**Sex-F1**) |
| IN -> IN | A (1,1) | 1.00 | 0.00 | 0.00 | 0.00 | 0.00 | -0.61 | 0.00 | 0.00 | 0.00 | 0.00 |
| IN -> ACC | A (2,1) | 1.00 | 0.00 | 0.00 | 0.98 | 0.78 | 0.48 | 0.00 | 0.00 | -0.20 | 0.07 |
| IN -> TH | A (3,1) | 1.00 | 0.99 | 0.00 | 0.00 | 0.00 | 0.26 | 0.17 | 0.00 | 0.00 | 0.00 |
| ACC -> IN | A (1,2) | 0.00 | 0.00 | 0.00 | 0.00 | 0.00 | 0.00 | 0.00 | 0.00 | 0.00 | 0.00 |
| ACC -> ACC | A (2,2) | 1.00 | 0.00 | 0.00 | 0.00 | 0.82 | -0.67 | 0.00 | 0.00 | 0.00 | 0.08 |
| ACC -> TH | A (3,2) | 1.00 | 0.00 | 0.96 | 0.94 | 0.00 | 0.11 | 0.00 | -0.28 | 0.29 | 0.00 |
| TH -> IN | A (1,3) | 0.00 | 0.00 | 0.51 | 0.00 | 0.00 | 0.00 | 0.00 | 0.06 | 0.00 | 0.00 |
| TH -> ACC | A (2,3) | 1.00 | 0.00 | 0.00 | 0.00 | 0.00 | -0.09 | 0.00 | 0.00 | 0.00 | 0.00 |
| TH -> TH | A (3,3) | 1.00 | 0.00 | 0.00 | 0.00 | 0.00 | -0.39 | 0.00 | 0.00 | 0.00 | 0.00 |
| **PEB: A matrix** | | **>=10% participants - 266** | | | | | | | | | |
| | | | P(**CP**) | P(**CP*EA**) | P(**CP*EN**) | P(**Sex-F1**) | E(**common**) | E(**CP**) | E(**CP*EA**) | E(**CP*EN**) | E(**Sex-F1**) |
| IN -> IN | A (1,1) | 1.00 | 0.00 | 0.00 | 0.00 | 0.00 | -0.64 | 0.00 | 0.00 | 0.00 | 0.00 |
| IN -> ACC | A (2,1) | 1.00 | 0.00 | 0.00 | 0.98 | 0.82 | 0.48 | 0.00 | 0.00 | -0.22 | 0.08 |
| IN -> TH | A (3,1) | 1.00 | 0.99 | 0.00 | 0.00 | 0.00 | 0.27 | 0.17 | 0.00 | 0.00 | 0.00 |

| | | | | | | | | | | | |
|---|---|---|---|---|---|---|---|---|---|---|---|
| ACC -> IN | A (1,2) | 0.00 | 0.00 | 0.54 | 0.00 | 0.00 | 0.00 | 0.00 | -0.07 | 0.00 | 0.00 |
| ACC -> ACC | A (2,2) | 1.00 | 0.00 | 0.00 | 0.00 | 0.91 | -0.70 | 0.00 | 0.00 | 0.00 | 0.11 |
| ACC -> TH | A (3,2) | 1.00 | 0.00 | 1.00 | 0.99 | 0.00 | 0.11 | 0.00 | -0.32 | 0.35 | 0.00 |
| TH -> IN | A (1,3) | 0.51 | 0.00 | 0.85 | 0.00 | 0.00 | -0.02 | 0.00 | 0.14 | 0.00 | 0.00 |
| TH -> ACC | A (2,3) | 1.00 | 0.00 | 0.00 | 0.00 | 0.00 | -0.10 | 0.00 | 0.00 | 0.00 | 0.00 |
| TH -> TH | A (3,3) | 1.00 | 0.00 | 0.00 | 0.00 | 0.00 | -0.40 | 0.00 | 0.00 | 0.00 | 0.00 |
| **PEB: A matrix** | | **>=15% participants - 176** | | | | | | | | | |
| | | | P(**CP**) | P(**CP*EA**) | P(**CP*EN**) | P(**Sex-F1**) | E(**common**) | E(**CP**) | E(**CP*EA**) | E(**CP*EN**) | E(**Sex-F1**) |
| IN -> IN | A (1,1) | 1.00 | 0.00 | 0.00 | 0.00 | 0.00 | -0.70 | 0.00 | 0.00 | 0.00 | 0.00 |
| IN -> ACC | A (2,1) | 1.00 | 0.00 | 0.00 | 0.91 | 0.71 | 0.50 | 0.00 | 0.00 | -0.24 | 0.07 |
| IN -> TH | A (3,1) | 1.00 | 0.72 | 0.00 | 0.00 | 0.00 | 0.24 | 0.10 | 0.00 | 0.00 | 0.00 |
| ACC -> IN | A (1,2) | 0.77 | 0.00 | 0.67 | 0.00 | 0.00 | 0.03 | 0.00 | -0.11 | 0.00 | 0.00 |
| ACC -> ACC | A (2,2) | 1.00 | 0.00 | 0.92 | 0.00 | 1.00 | -0.74 | 0.00 | -0.22 | 0.00 | 0.14 |
| ACC -> TH | A (3,2) | 1.00 | 0.00 | 1.00 | 0.92 | 0.00 | 0.12 | 0.00 | -0.32 | 0.31 | 0.00 |
| TH -> IN | A (1,3) | 1.00 | 0.00 | 1.00 | 0.00 | 0.00 | -0.06 | 0.00 | 0.27 | 0.00 | 0.00 |
| TH -> ACC | A (2,3) | 1.00 | 0.00 | 0.87 | 0.00 | 0.00 | -0.13 | 0.00 | -0.19 | 0.00 | 0.00 |
| TH -> TH | A (3,3) | 1.00 | 0.00 | 0.00 | 0.00 | 0.00 | -0.42 | 0.00 | 0.00 | 0.00 | 0.00 |

*Table 15* ***BMA results of intrinsic connections using a PEB matrix with multiple covariates, CP*EA (Chronic pain and history of emotional abuse), CP*EN (Chronic pain and history of emotional neglect) and SEX-F1 (female coded as 1), using only data acquired in Dundee****. P(x) shows the posterior probability of the estimates (E(x)) being non-zero. The first column identifies the connection as A(TO,FROM), where 1=insula, 2=anterior cingulate cortex, 3=thalamus; e.g. A(1,2) is the connection from ACC to insula.*
